# Clinical characteristics of the recovered COVID-19 patients with re-detectable positive RNA test

**DOI:** 10.1101/2020.03.26.20044222

**Authors:** Jianghong An, Xuejiao Liao, Tongyang Xiao, Shen Qian, Jing Yuan, Haocheng Ye, Furong Qi, Chengguang Shen, Yang Liu, Lifei Wang, Xiaoya Cheng, Na Li, Qingxian Cai, Fang Wang, Jun Chen, Yingxia Liu, Yunfang Wang, Feng Zhang, Yang Fu, Xiaohua Tan, Lei Liu, Zheng Zhang

## Abstract

**Background:** It has been reported that several cases recovered from COVID-19 tested positive for SARS-CoV-2 after discharge (re-detectable positive, RP), however the clinical characteristics, significance and potential cause of RP patients remained elusive.

**Methods:** A total of 262 COVID-19 patients were discharged from January 23 to February 25, 2020, and were enrolled for analysis of their clinical parameters. The RP and non-RP (NRP) patients were grouped according to the disease severity during their hospitalization period. The clinical characterization at re-admission to the hospital was analyzed. SARS-CoV-2 RNA and plasma antibody levels were detected using high-sensitive detection methods.

**Findings:** Up to March 10, 2020, all of patients were followed up for at least 14 days, and 38/262 of RP patients (14.5%) were present. The RP patients were characterized by being less than 14-years old and having mild and moderate conditions as compared to NRP patients, while no severe patients became RP. Retrospectively, the RP patients displayed fewer symptoms, more sustained remission of CT imaging and earlier RNA negative-conversion but similar plasma antibody levels during their hospitalization period as compared to those NRP patients. When re-admitted to the hospital, these RP patients showed no obvious clinical symptoms or disease progression indicated by normal or improving CT imaging and inflammatory cytokine levels. All 21 close contacts of RP patients were tested negative for SARS-CoV-2 RNA, and no suspicious clinical symptoms were reported. However, 18/24 of RNA-negative samples detected by the commercial kit were tested to be positive for virus RNA using a hyper-sensitive method, suggesting the carrier status of virus possibly existed in patients recovered from COVID-19.

**Interpretation:** Our results showed that young and mild COVID-19 patients seem to be RP patients after discharge, who show no obviously clinical symptoms and disease progression upon re-admission. More sensitive RNA detection methods are required to monitor these patients during follow-up. Our findings provide empirical information and evidence for the effective management of COVID-19 patients during their convalescent phase.

## Introduction

Since early December of 2019 and up to March 23, 2020, over 300, 000 cases of coronavirus disease 2019 (COVID-19) caused by novel coronavirus (SARS-CoV-2) infection, with over 13, 000 deaths have been reported through the world [1]. The World Health Organization has declared COVID-19 as a pandemic [2]. Generally, the COVID-19 is less severe and less fatal than the SARS, however, some patients, especially those who are elderly with co-morbidities are prone to develop more severe symptoms and require emergent medical interventions [3, 4]. Many literatures have retrospectively analyzed the clinical characteristics of patients infected with SARS-CoV-2 [3-8]. Recently, an increasing number of patients with COVID-19 were discharged from the hospital and received regular follow-up and observation. Re-detectable positive (RP) of SARS-CoV-2 RNA test in some recovered patients has been reported [9-12]. The management of RP patients has attracted wide attention. However, the number of RP patients reported in the literature was small, and the duration of follow-up was short. In addition, the clinical characteristics is lacking and the potential impact and significance of RP patients remain unknown, which makes it difficult to provide empirical information and evidence support for the management of patients with COVID-19 in the recovery period.

This study retrospectively analyzed the clinical characteristics of 38 RP patients and 224 non-RP (NRP) patients recovered from COVID-19. It is found that RP patients were characterized by younger age and milder conditions. They also had minor symptoms, more sustained remission of CT imaging and earlier RNA negative-conversion but similar plasma antibody levels during their hospitalization period. They showed no obvious disease progression and infectivity when re-admitted to the hospital. The hyper-sensitive detection method identified SARS-CoV-2 RNA molecules from most samples that were tested RNA-negative by the commercial kit, suggesting the carrier status of virus possibly existed in recovering COVID-19 patients. These findings provide key information for the effective management of COVID-19 patients during their convalescent phase.

## Methods

### Study design and participants

A total of 262 confirmed COVID-19 patients discharged from Shenzhen Third People’s Hospital from January 23, 2020 to February 25, 2020 were enrolled in this study. All discharged COVID-19 patients were continued to be isolated and observed for 14 days, weekly followed-up and SARS-CoV-2 RNA detection were performed timely. All discharged patients were followed-up for at least two additional weeks after isolation. Among them, the RP patients were re-admitted to hospital for further medical observation and close contacts were also followed-up. The rest of the recovered NRP patients were closely followed-up outside the hospital. This study was approved by the Ethics Committee of The Third People’s Hospital of Shenzhen (2020-115), which waived the requirement for written patient consent for this retrospective analysis. All patients gave their oral consent to participate in this retrospective study.

### Clinical definition

According to the guideline of the diagnosis and treatment for novel coronavirus pneumonia (the sixth edition) published by National Health Commission of the People’s Republic of China [13], all first diagnosis cases of COVID-19 were confirmed according to positive respiratory RT-PCR tests. The discharge criteria of the recovered patients included: temperature returned to normal for more than 3 days, respiratory symptoms significantly improved, and significant absorption of pulmonary lesions of chest CT imaging, and at least consecutive negative RNA test results for 2 apart from each other by at least 24 hours. The RP patients were confirmed by digestive (anal swab) and respiratory positive RT-PCR tests. Since February 22, 2020, evaluation of negative anal swab was supplemented for discharge criteria in Shenzhen Third People’s Hospital.

### Data collection

The medical records of 262 recovered COVID 19 patients including 38 RP patients were reviewed. The epidemiological, demographic, clinical, laboratory data of the patients were collected, summarized and analyzed. According to the first chest CT imaging post admission, the extent of pulmonary inflammation was divided into mild, moderate and severe condition basing on the lesions involving unilateral lobe, multiple lobes in both lungs, and all lobes in both lungs, respectively. According to chest CT within 7 days after admission, the remission of the lesions was evaluated. The temporary progression was indicated by increased lesion and persistent remission was indicated by stable or absorbed or decreased lesions.

### qRT-PCR and Sherlock assay for SARS-CoV-2 RNA detection

The quantitative reverse transcription polymerase chain reaction (qRT-PCR) was assessed as described previously [14]. Nasopharyngeal and anal specimens collected during hospitalization were sent to the laboratory in viral transport case. Total nucleic acid extraction were extracted from the samples using the QIAamp RNA Viral Kit (Qiagen, Heiden, Germany), and quantitative RT-PCR was performed using a China Food and Drug Administration (CFDA) approved commercial kit specific for 2019-nCoV detection (GeneoDX Co., Ltd., Shanghai, China) or Sherlock kit gifted from Feng Zhang lab according to the manual. Each RT-PCR assay provided a Ct value, which is the number of cycles required for the fluorescent signal to cross the threshold for a positive test, a higher Ct value is correlated with a lower viral load. The specimens were considered positive if the Ct value was ≤ 37.0, and negative if the viral load were undetectable. Specimens with a cycle-threshold value higher than 37 were repeated. The specimen was considered positive if the repeat results were the same as the initial result and between 37 and 40. If the repeat Ct was undetectable, the specimen was considered negative. All procedures involving clinical specimens and SARS-CoV-2 were performed in a biosafety level 3 laboratory. Meanwhile, we did next-generation sequencing of samples from three patients.

### ELISA assay for anti-SARS-CoV-2 IgG and IgM antibody

Microtiter plates (Sangon Biotech) were coated overnight at 4°C with 4 μg/mL recombinant SARS-CoV-2-RBD (Receptor binding domain) proteins (50 μL per well) expressed by our laboratory through 293-T cells. The plates were washed thrice with PBS containing 0.1% v/v Tween-20 (PBST) and blocked with blocking solution (PBS containing 2% w/v non-fat dry milk) for 2 hours at 37°C. The plates were then washed with PBST. The sera were diluted to 200-fold into PBS as an initial concentration, and serial 3-fold dilutions of sera was added to the wells and incubated at 37°C for 60 minutes. After three washes, 100 μL of horseradish peroxidase (HRP)-conjugated goat anti-human IgG (for IgG antibody titer detection) and IgM (for IgM antibody titer detection) antibodies solution (Sangon Biotech) were added to each plate, respectively, and incubated at 37°C for 60 minutes. After five washes, 100 μL of tetramethylbenzidine (TMB) substrate (Sangon Biotech) was added at room temperature in the dark. After 15 minutes, the reaction was stopped with a 2 M H2SO4 solution. The absorbance was measured at 450 nm. All samples were run in triplicate. The ELISA titers were determined by endpoint dilution.

### Statistical analysis

A statistical analysis was performed using SPSS 26.0 (IBM, Chicago). All of the statistical tests were two-sided, and significant differences were considered at p < 0.05. Continuous variables were evaluated using the median and interquartile range (IQR) values. Chi-square or Fisher exact tests were utilized to compare the proportions of the categorical variables.

## Results

### Demographic, epidemiological and clinical characteristics

A total of 262 patients were discharged from January 23, 2020 to February 25, 2020 and were followed-up for at least 14 days. Among them, mild, moderate and severe patients accounted for 11.4% (n = 30), 81.0% (n = 212) and 7.6% (n = 20), respectively. Up to March 10, 14.5% of convalescent patients (n = 38) were re-detected to be SARS-CoV-2 RNA positive during their followed-up period. None of severe patients were re-tested to be RNA positive (Supplemental Table 1).

**Table 1.** Baseline characteristics of enrolled patients with COVID-19.

|  | Mild (n = 30) |  |  | Moderate (n = 212) |  |  |
| --- | --- | --- | --- | --- | --- | --- |
|  | RP (n = 11) | NRP (n = 19) | *P*<br>value | RP (n = 27) | NRP (n = 185) | *P*<br>value |
| Age, median (IQR)-yr | 20 (5-64) | 23 (2-63) | 0.98 | 38 (2-60) | 48 (1-86) | < 0.01 |
| > 14 and ≤ 60 years old-n.(%) | 10 (90.9) | 18 (94.7) | 0.78 | 26 (96.3) | 139 (75.1) | 0.11 |
| ≤ 14 years old-n.(%) | 4 (36.3) | 6 (31.6) | 0.57 | 3 (11.1) | 6 (3.24) | 0.04 |
| > 60 years old-n.(%) | 1 (9.1) | 1 (5.3) | 0.32 | 1 (3.7) | 46 (24.9) | < 0.001 |
| Gender-n.(%) |  |  |  |  |  |  |
| Male | 4 (36.3) | 10 (52.6) | 0.08 | 12 (44.4) | 90 (48.6) | 0.66 |
| Female | 7 (63.7) | 9 (47.4) | 0.12 | 15 (55.6) | 95 (51.4) | 0.69 |
| Comorbidities-n.(%) | 1 (9.1) | 0 (0) | NA | 1 (3.7) | 41 (22.2) | < 0.01 |
| History of travel or residence in Hubei-n.(%) | 10 (90.9) | 16 (84.2) | 0.61 | 23 (85.2) | 152 (82.2) | 0.82 |
| Fever--n.(%) | 2 (18.2) | 7 (36.8) | < 0.01 | 23 (85.2) | 133 (71.9) | 0.29 |
| Upper respiratory symptoms-n.(%) | 5 (45.5) | 2 (10.5) | < 0.01 | 4 (14.8) | 34 (18.4) | 0.53 |
| Lower respiratory symptoms-n.(%) | 5 (45.5) | 7 (36.8) | 0.34 | 14 (51.9) | 95 (51.4) | 0.96 |
| Digestive tract symptoms-n.(%) | 0 (0) | 2 (10.5) | NA | 3 (11.1) | 15 (8.11) | 0.50 |
| The lesion range of chest CT-n.(%) |  |  |  |  |  |  |
| Unilateral | 0 (0) | 0 (0) | NA | 6 (22.2) | 36 (19.4) | 0.66 |
| Multi-lobe of Bilateral | 0 (0) | 0 (0) | NA | 15 (55.6) | 105 (56.7) | 0.92 |
| All-lobe of Bilateral | 0 (0) | 0 (0) | NA | 6 (22.2) | 44 (23.7) | 0.82 |
| Chest CT imaging-n(%) |  |  |  |  |  |  |
| Transient progression | 0 (0) | 0 (0) | NA | 4 (14.8) | 67 (36.2) | < 0.05 |
| Sustained remission | 0 (0) | 0 (0) | NA | 23 (85.2) | 113 (61.1) | 0.05 |
| Steroids use-n.(%) | 0 (0) | 0 (0) | NA | 4 (14.8) | 27 (14.6) | 0.97 |

RP, re-detectable positive patients; NRP, non-re-detectable positive patients

It revealed that the vast majority of RP patients (97.4%, n = 37) were younger than 60 years of age. Among them, patients younger than 14 years old were more common compared with those between the ages of 14 and 60 years (35.0% vs 16.0%, p < 0.01) (Table 1). In addition, it is found that 36.7% (11/38) of RP patients are characterized by mild symptoms. The percentage was significantly higher than what was seen among NRP patients (12.7%, 19/204, p < 0.01, Supplemental Table 1). There was no significant difference in the gender distribution. Notably, there were less mild RP patients having fever in their initial symptoms as compared to mild NRP patients (p < 0.01). Also, 45.5% of mild RP patients displayed only upper respiratory symptoms at the first admission, while mild NRP patients usually had lower respiratory symptoms at the first admission (Table 1). There is no difference of the extent of lesions in the first chest CT imaging between RP and NRP patients with moderate stages. However, the incidence of RP (85.2%) was found to be particularly closely related to the sustained remission of chest CT imaging as compared to NRP patients, of which 36.2% displayed transient progression during their first hospitalization period (Table 1 and Figure 1). There was no significant difference in the usage of steroid and antiviral therapy between RP and NRP patients during their first hospitalization period. In addition, RP patients did not show a higher incidence of a history of traveling and living in Hubei province as compared to NRP patients.

**Figure 1.**
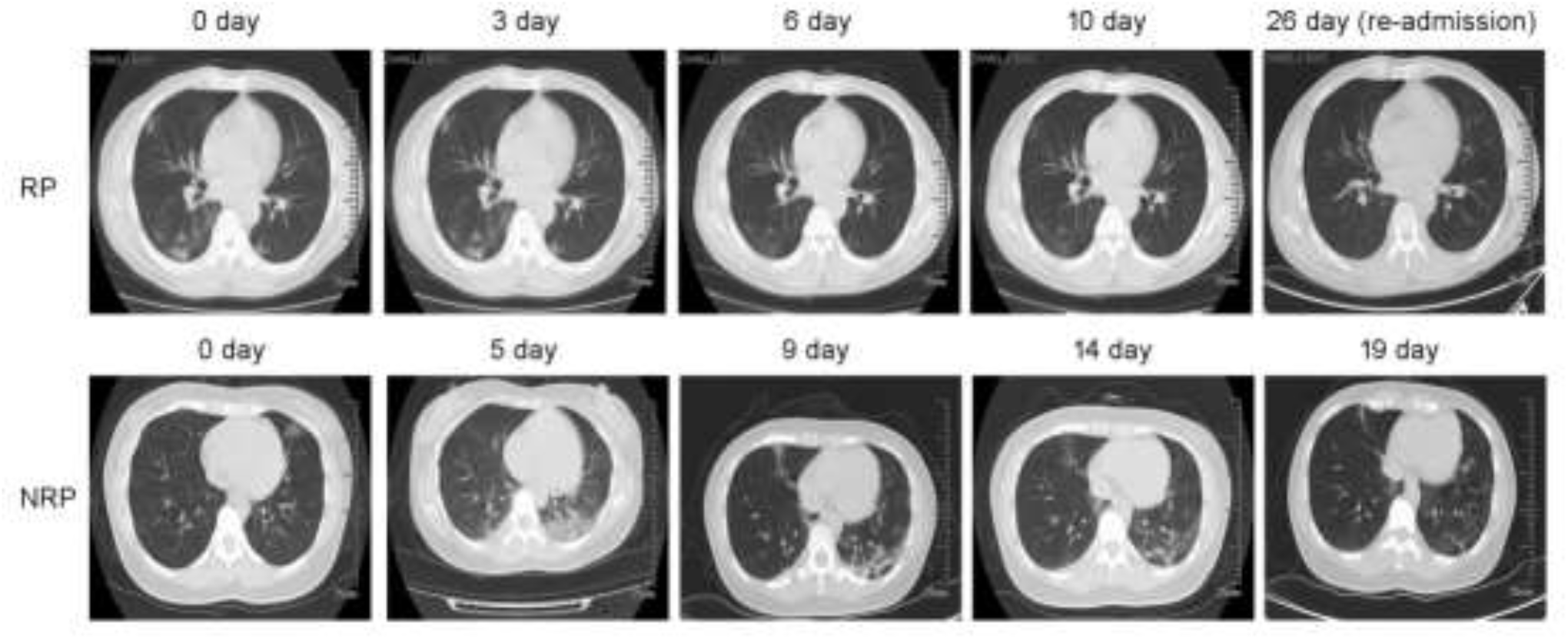
Serial CT imaging of a representative RP and NRP patient. For the RP patient, the first chest CT scan on admission (day 7 since the onset of illness) showed ground-glass opacity in both lungs. At 3 day, 6 day and 10 day after admission (day 10, day 13 and day 17 since the onset of illness), the lung lesions on the chest CT imaging was significantly reduced accompanied by the disappeared clinical symptoms. The patient was discharged at day 12 after admission (day 19 since the onset of illness). At day 26 (day 33 since the onset of illness), the patient was re-admitted without fever and cough due to positive RNA detection. The chest CT showed no inflammatory lesions. For the NRP patient, a chest CT scan showed a small ground glass in the upper left lung on admission (day 3 since the onset of illness). On day 2 and 8 after admission (day 5 and day 11 since the onset of illness), the double lower lung lesions increased significantly on chest CT imaging although the body temperature and the oxygenation index was returned to normal levels. On day 9, 14 and 17 after admission (day 12, day 17 and day 20 since the onset of illness), the lesions in both lower lungs were recovered on chest CT imaging. Then the patient was discharged without fever and cough at day 18 after admission (day 21 since the onset of illness) when SRAS-CoV-2 RNA was also detected to be negative.

### Differential RNA dynamic in RP patients with that in NRP patients

No differences of days of last RNA negative-conversion since the onset of illness and hospitalization days were found between RP and NRP patients. Importantly, RNA negative-conversion occurred mostly within 2-3 weeks since the onset of illness among 63.6% mild RP patients and within 1-2 weeks since the onset of illness among 22.2% moderate RP patients. By contrast, there are more NRP patients who displayed RNA negative-conversion after 3 weeks since the onset of illness regardless of mild or moderate status (Table 2). These data showed RP patients were characterized by early RNA-negative conversion while NRP patients cleared virus relatively late.

**Table 2.**
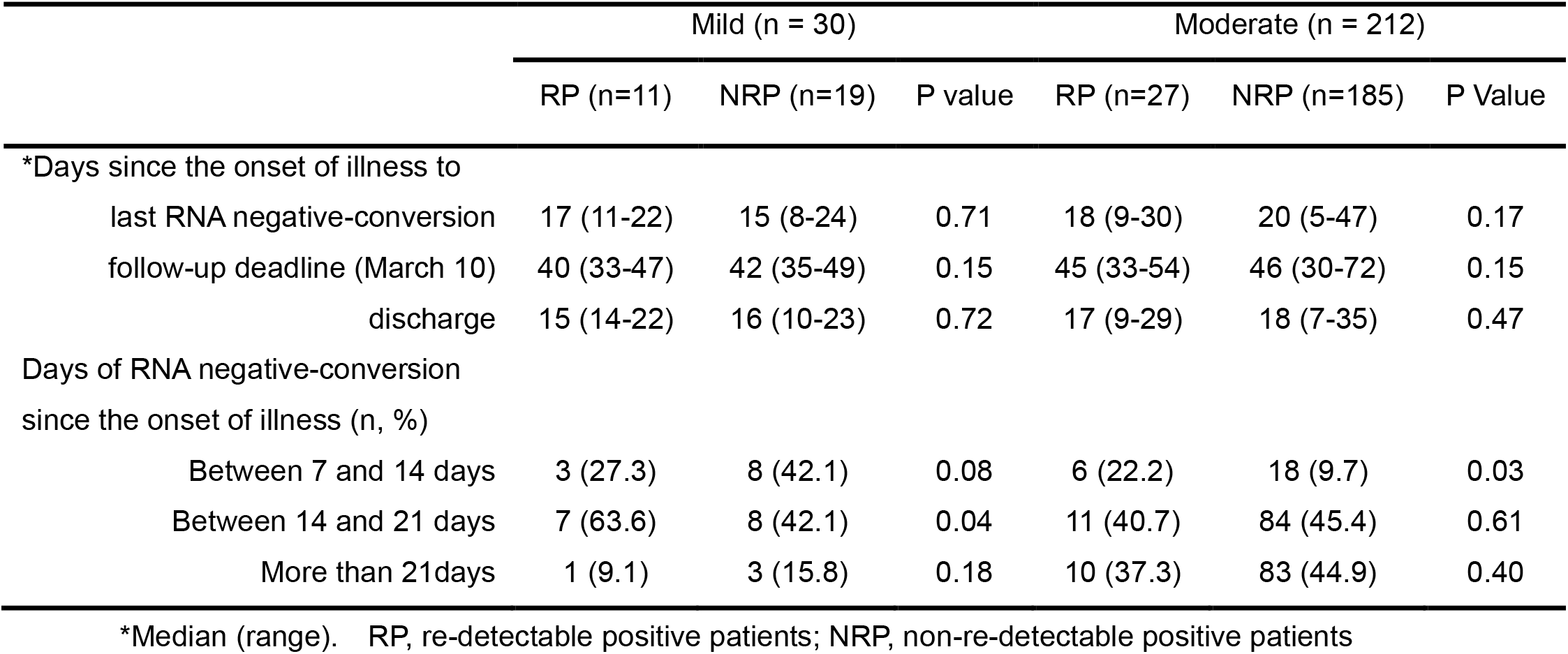
RNA detection in the enrolled patients with COVID-19.

### The changes of serum anti-SARS-CoV-2 IgG and IgM antibodies in patients recovered from COVID-19

In order to evaluate the effect of serum-specific antibody levels on the occurrence of RP, we analyzed the difference of anti-SARS-CoV-2 IgG and IgM antibody levels in both RP and NRP patients at their discharge. More than half of RP and NRP patients displayed medium levels of IgG and IgM independent on their disease severity. However, there are no differences of antibody levels in between both groups of patients (Supplemental Table 2). We also evaluated the dynamic of IgG and IgM levels at the discharge and re-admission in RP patients. The IgG and IgM levels were maintained at stable levels in these RP patients during the 14 days period (data not shown).

### Supplementing negative results of anal swab test at discharge failed to reduce RP occurrence of COVID-19 patients

In order to evaluate the effects of increasing sampling site test at discharge on RP events, we compared the occurrence events of RP before Feb 22 and after Feb 22 after that time the negative anal swab detection was added to discharge criterion in COVID-19 patients. Our results showed that there was no statistical difference in the occurrence of RP patients before Feb 22 and after Feb 22 (14.5% *vs* 14.3%, *p* = 0.77, Figure 2). These data indicated that supplementing detected sites of SARS-CoV-2 RNA failed to reduce the RP occurrence in convalescent patients.

**Figure 2.**
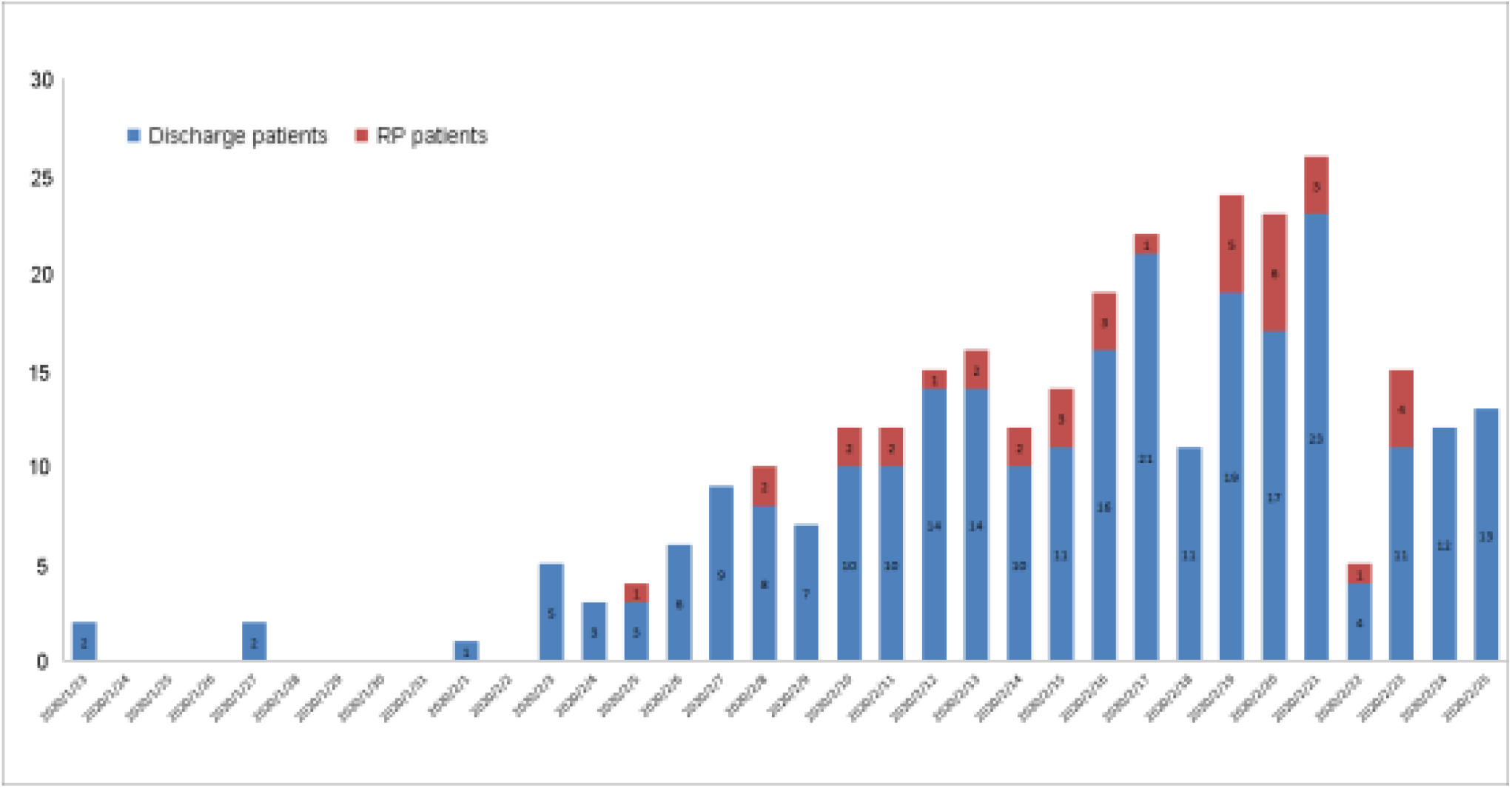
The number of discharge patients and RP patients each day from Jan 23 to March 10, 2020. On Feb 22, 2020, the anal swab negative test was added to discharge criterion. Blue, the number of discharge patients. Red, the number of RP patients.

### RP patients showed no obvious clinical symptoms and disease progression

All 38 RP patients were re-admitted to the hospital for further medical observation. The analysis showed that all these patients had no fever. A small number of patients reported mild cough and chest tightness, which were not worse than before (Table 3). All patients recovered from mild conditions (n = 30) and 37.0% from moderate patients had normal chest CT imaging without inflammatory signs. By contrast, 63.0% (n = 17) of patients recovered from moderate conditions had stable or reduced inflammatory signs in their chest CT imaging (Figure 1). There were normal range of the lymphocyte count, plasma IL-6 and CRP levels upon admission for all RP patients. Only one patient received transient interferon-alpha inhalation therapy, and 4 patients received low-flow oxygen inhalation therapy and traditional Chinese medicine after admission.

**Table 3.** Clinical observation of RP patients at re-admission of hospital.

|  | Mild (n = 11) | Moderate (n = 27) |
| --- | --- | --- |
| Symptoms |  |  |
| fever | 0 (0) | 0 (0) |
| cough | 1 (9.1) | 5 (18.5) |
| Chest tightness | 0 (0) | 2 (7.4) |
| other | 0 (0) | 3 (11.1) |
| Chest CT imaging |  |  |
| Normal | 11 (100) | 10 (37.0) |
| Stable or absorb | 0 (0) | 17 (63.0) |
| Progression | 0 (0) | 0 (0) |
| Laboratory examination |  |  |
| Abnormal lymphocyte count | 0 (0) | 4 (14.8) |
| increasing serum IL-6 level | 0 (0) | 0 (0) |
| increasing serum CRP level | 0 (0) | 0 (0) |
| Treatment |  |  |
| Low flow oxygen | 0 (0) | 4 (14.8) |
| Traditional Chinese medicine | 3 (27.3) | 8 (29.6) |
| Antiviral therapy | 0 (0) | 1 (3.7) |
| Number of contacts with symptoms | 0 | 0 |

**Table 4.** The comparison between hyper-sensitivity and common sensitivity detection in RP and NRP patients.

|  | Sample | Dates since the onset of illness | Sherlock |  | Commercial |  |
| --- | --- | --- | --- | --- | --- | --- |
|  |  |  | S gene | ORF gene | N gene | ORF1 gene |
| 1 | Anal swab | 44 | - | - | - | - |
| 2 | Nasal swab | 44 | + | - | - | - |
| 3 | Anal swab | 44 | + | - | - | - |
| 4 | Nasal swab | 44 | + | + | - | - |
| 5 | Anal swab | 37 | - | + | + | - |
| 6 | Nasal swab | 37 | + | + | - | - |
| 7 | Anal swab | 42 | + | - | - | - |
| 8 | Anal swab | 43 | - | + | - | - |
| 9 | Anal swab | 30 | + | + | - | - |
| 10 | Blood | 42 | + | - | - | - |
| 11 | Blood | 43 | - | - | - | - |
| 12 | Blood | 30 | + | - | - | - |
| 13 | Nasal swab | 42 | - | - | + | - |
| 14 | Nasal swab | 43 | + | - | - | - |
| 15 | Nasal swab | 30 | + | + | - | - |
| 16 | Anal swab | 32 | + | + | - | - |
| 17 | Anal swab | 37 | + | - | - | - |
| 18 | Anal swab | 36 | + | - | - | - |
| 19 | Anal swab | 37 | + | + | - | - |
| 20 | Anal swab | 41 | + | + | - | - |
| 21 | Anal swab | 37 | - | - | + | + |
| 22 | Anal swab | 31 | + | - | - | - |
| 23 | Anal swab | 43 | + | + | - | - |
| 24 | Nasal swab | 32 | + | - | - | - |
| total | Positive (%) |  | 18 (75%) | 10 (41.6%) | 3 (12.5%) | 1 (4.2%) |
| 25 | Nasal swab | 47 | - | - | - | - |
| 26 | Anal swab | 47 | - | - | - | - |
| 27 | Nasal swab | 45 | - | - | - | - |
| 28 | Nasal swab | 40 | - | - | - | - |
| 29 | Anal swab | 54 | - | - | - | - |
| 30 | Anal swab | 49 | - | - | - | - |
| 31 | Nasal swab | 44 | - | - | - | - |
| 32 | Nasal swab | 48 | - | - | - | - |
| total | Positive (%) |  | 8 (0) | 8 (0) | 8 (0) | 8 (0) |

Note: 1-24, re-detectable positive patients; 25-32: non-re-detectable positive patients.

In addition, because all the convalescent patients with COVID-19 in our cohort were required to be isolated at home or under intensive isolation, only 21 close contacts were produced. Up to March 10, 2020, all of 21 close contacts were tested to be negative for SARS-CoV-2 RNA, and no suspicious clinical symptoms were reported in those close contacts.

### Hyper-sensitive methods potentially improved SARS-CoV-2 RNA detection in RP patients

To investigate the possible false negative due to low-sensitive commercial RNA detection kit, we used a higher-sensitive method to detect various types of samples from both RP patients and NRP patients with similar illness days. For 24 samples from 15 of RP patients who were sampled after 5-7 days since the onset of the re-admission, 75% of spike genes and 41.6% ORF genes could be detected to be positive using hyper-sensitive method, while only 12.5% N genes and 4.2% RF genes were detected to be positive using commercial detection kit. Eight of fifteen RP patients were confirmed to be RNA positive using the hyper-sensitive kit, although only 1 person was confirmed using commercial kit. By contrasts, 8 samples from NRP patients were detected to be negative by both methods. These data showed hyper-sensitive methods potentially improve RNA positive detection in samples from RP patients with negative results.

## Discussion

Several studies have shown the existence of RP patients [9-12], however their clinical characterization as not well defined. This study retrospective analyzed the clinical and followed-up data in a cohort of RP and NRP patients in the same discharge period. Up to March 10, 2020, 38 RP patients were present which accounts for 14.5% of discharged patients during the same followed-up period. These RP patients displayed several significant features, including younger age and mild and/or moderate symptom during their hospitalization, which is in consistent with a previous report [9, 12]. Mild RP patients were usually younger than 14 years old and moderate RP patients were younger than 60 years old. By contrast, no severe patients were found to be RP patients within the similar follow-up period. In addition, more RP patients displayed minor symptoms in their hospitalization such as less comorbidities and fever, and more upper respiratory symptoms. RP patients also maintained more remission in their CT imaging than those of NRP patients. These data indicated that RP patients were characterized by younger and minor symptoms in their hospitalization period.

Virus load is usually thought to be related to the disease outcome [14, 15]. The present study indicated RNA negative-conversion occurred commonly 2-3 weeks since the onset of illness in moderate RP patients as compared to more than 3 weeks in moderate NRP patients. The significantly shortened RNA negative-conversion time may affect the persistence of high levels of adaptive immunity [16]. Our recent studies indicated that a higher titer of antibody in the plasma was independently associated with disease severity in patients with COVID-19 [17]. However, RP and NRP patients displayed similar levels of IgG and IgM in the plasma. Future study should investigate host immune responses which were usually considered to determine the clinical outcome especially in virus infection [18, 19].

We also comprehensively characterized the clinical symptoms of RP patients when they were re-admitted to the hospital. No obvious clinical evidence of disease progression or recurrence was found in these RP patients including CT imaging and laboratory tests. And no antibiotics, steroids, antiviral agents and continuous supplemental oxygenation were required in these RP patients. The inflammatory response was significantly reduced. These data indicated that the diseases of RP patients did not progress into more severe status even if their RNA was detected to be positive for SARS-CoV-2. More important, these RP patients have not caused new infections after discharge. And from a recent longitudinal study in SARS-CoV-2 infected rhesus macaques, reinfection could not occur in convalescent monkeys [20]. Long-term follow-up of these close contacts with RP patients will warrant the evaluation of possible risk of RP.

The underlying mechanisms underlying RP occurrence remain unclear. The possible reasons argued by a large number of experts are related to several virological, immunological and sampling methodological factors. Virologically, the false negatives [21], viral residual [12], intermittent viral release [12] and viral distribution [22, 23] are usually considered to be major factors. Our data support the notion that the false negatives using commercial kit may partially account for the RP, because the kits had only 30%–50% positive rate of detection [23, 24]. In 24 of various samples from RP patients, RNA was detected to be negative for both N gene and ORF1b gene at several days after their re-admission to the hospital using commercial kit, whose lower limit of detection (LOD) was relatively high (500 copies/ml). However, using a more sensitivity Sherlock kit with an LOD of 100 copies/ml [25], 75% of samples were detected to be positive for S gene and 41.6% for ORF genes, thus leading to half of positive subjects present within RP patients with undetectable RNA using commercial kit in their hospitalization. By contrast, among 8 samples from NRP patients, none was detected to be positive using either Sherlock or commercial kit. However, in a sample confirmed by SRAS-CoV-2 sequencing, the Sherlock tested it as positive (data not shown). Therefore, future study should improve both the sensitivity and specificity of detection kit, which would accurately identified clinical samples. Anyway, these data indicated that false positive detected by current kit may on some extent account for the occurrence of RP patients.

Another virological factor is that long-term virus residual in gut and other tissue, similar to SARS [26]. A recent study indicated that SARS-CoV-2 nucleic acid can persist in the digestive tract and feces for nearly 50 days [27]. Thus, extending detection time is necessary for the COVID-19 patients when they were discharged. However, our results show that adding anal swab derived RNA tested to be negative as discharge criterion did not significantly reduce the occurrence of RP patients. Thus other factor may be associated with the RP patients. We could not exclude sampling methodological factors including differential sampling and operational methods, sample quality, and technician expertise levels. Nor could we exclude immunological factors including low mucosal immune responses such as low IgA levels. These factors may take some uncertain risks leading the occurrence of RP patients [4, 27]. Future studies should reduce RP occurrence through using hyper-sensitive detection kit with hyper-specificity, combining detection of multiple samples with more immune markers.

This study has several limitations. First, this study is a single-center retrospective study and the duration of follow-up is short, and more clinical observations are needed to evaluate the potential risk of SARS-CoV-2 recurrence and infection. Second, dynamics of SARS-CoV-2 RNA in COVID-19 patients need to be monitored and evaluated for RP patients. Second, additional studies should measure the dynamic changes of serum specific antibody levels in RP patients and evaluate the continuous protective effect of serum specific antibodies on patients with COVID-19. Finally, we should differentiate RP patients from relapse ones from convalescent subjects, for who two distinct prevention and control strategies will be adopted.

Taken together, our findings revealed the clinical features of RP patients who did not show recurrence of clinical symptoms and abnormal laboratory tests. However, hyper-sensitive detection methods revealed the existence of SARS-CoV-2 RNA in RP patient specimens tested to be negative using the commercial kit. Therefore, it is necessary to develop a more accurate quantitative assessment of the RNA dynamics and additional discharge criteria to help physicians make a decision. This study provided valuable empirical information and clinical evidence support for effective management of COVID-19 patients during convalescent period. Further study should evaluate the potential clinical significance and transmission risk of RP patients.

## Contributors

ZZ, LL, XT, and JA had the idea and for designed the study. ZZ, LL, XT, JA, XJ and QS had full access to all data in the study and take responsibility for the integrity of the data and the accuracy of the data analysis. JY, LW, XC, SQ, NL, QC, FW, JC, YL, LL and ZZ had roles in the clinical management, patient recruitment, sample preparation and clinical data collection. TX and CS had roles in the RNA and antibody detection experiments, data collection and analysis. JA, HY, FQ, YL and ZZ had roles in statistical analysis. JA, XJ, YW, FZ, XT, LL, YL and ZZ had roles in data interpretation. JA, XL, YF, XT and ZZ wrote the manuscript. YW, YF, XT and ZZ contributed to critical revision of the report. All authors reviewed and approved the final version of the manuscript.

## Data Availability

I have full access to all the data in the study and had final responsibility for the decision to submit for publication.

## Acknowledgements

We acknowledge the work and contribution of all the health providers from Shenzhen Third People’s Hospital.

## Research in context

### Evidence before this study

Positive SARS-CoV-2 RNA test in some recovered patients has been reported, whose management has attracted wide attention. However, the number of recovered patients with the positive SARS-CoV-2 RNA test reported in the literature was small, and the duration of follow-up was short. The clinical characteristics are lacking and the potential impact and significance of the patients remain unknown. The lack of these data makes it difficult to provide empirical information and evidence support for the management of patients with COVID-19 in the recovery period.

### Added value of this study

First, the young and mild COVID-19 patients are prone to be tested positive for SARS-CoV-2 after discharge. These patients display fewer symptoms, more sustained remission of CT imaging and earlier RNA negative-conversion but similar plasma antibody levels during the hospitalization period as compared to those NRP patients. Second, upon re-admission, these patients show no obviously clinical symptoms and disease progression. However, the hyper-sensitive detection method potentially recognized false negative by the commercial kit.

### Implications of all the available evidence

The current evidence strongly supports the effective management of COVID-19 patients during their convalescent phase.

cases recovered from COVID-19 tested positive for SARS-CoV-2 after discharge (re-detectable positive, RP)

**Supplemental Table 1.** Analysis of disease severity in RP and NRP patients.

|  | Mild (n = 30) | Moderate (n = 212) | Severe (n = 20) | Total (n = 262) |
| --- | --- | --- | --- | --- |
| RP | 11 (36.7%) | 27 (12.7%) | 0 (0) | 38 (14.5%) |
| NRP | 19 (63.3%) | 185 (87.3%) | 20 (100%) | 224 (85.5%) |

RP, re-detectable positive patients; NRP, non-re-detectable positive patients

**Supplemental Table 2.** Analysis of serum anti- SARS-CoV-2 IgG and IgM antibody levels in patients with covid-19 at discharge

|  | Mild |  |  | Moderate |  |  |
| --- | --- | --- | --- | --- | --- | --- |
|  | RP (n = 10) | NRP (n = 10) | P value | RP (n = 21) | NRP (n = 119) | P value |
| Serum IgG (n, %) |  |  |  |  |  |  |
| High | 1 (10%) | 0 (0) | 0.30 | 8 (38.1%) | 41 (34.5%) | 0.95 |
| Medium | 6 (60%) | 4 (40.0%) |  | 10 (47.6%) | 60 (50.4%) |  |
| Low | 3 (30%) | 6 (60.0%) |  | 3 (14.3%) | 18 (15.1%) |  |
| Serum IgM (n, %) |  |  |  |  |  |  |
| High | 1 (10%) | 0 (0) | 0.59 | 7 (33.3%) | 39 (32.8%) | 0.68 |
| Medium | 7 (70%) | 8 (80.0%) |  | 12 (57.1%) | 60 (50.4%) |  |
| Low | 2 20%) | 2 (20.0%) |  | 2 (9.5%) | 20 (16.8%) |  |

Note: RP, re-detectable positive patients; NRP, non-re-detectable positive patients
High, titer with more than 16400; Medium, titer between 5400 and 16400; Low, titer with less than 5400

